# Polygenic Determinants of Arterial Stiffness: Implications for Hypertension

**DOI:** 10.1101/2025.10.21.25338503

**Authors:** Adam D. Gepner, Ryan J. Pewowaruk, Claudia E. Korcarz, Adriana Hung, Kent D. Taylor, Y.-D. Ida Chen, Alexis C. Wood, Jie Yao, Xiuqing Guo, Jerome I. Rotter

## Abstract

**Background:** Components of arterial stiffness (structural stiffness from aging and load-dependent stiffness from high blood pressure) are markers for cardiovascular disease risk. We evaluated relationships between polygenic risk scores (PRS) for blood pressure (BP) and arterial stiffness.

**Methods:** Participants (n=5942) are from the Multi-Ethnic Study of Atherosclerosis with measures of carotid ultrasound and genomic DNA. Pulse wave velocity (PWV) was calculated from carotid ultrasound representing total stiffness. Structural stiffness was calculated by adjusting PWV to a 120/80 mmHg blood pressure with participant-specific models. Load-dependent stiffness was the difference of total and structural stiffness. Blood pressure PRSs were previously derived from a genome-wide association study. Associations between BP PRSs and components of carotid arterial stiffness were assessed with multivariable-adjusted regression models. Mendelian Randomization (MR) was performed with two-stage least squares to 1) confirm that BP PRSs strongly predict MAP and 2) estimate the effect of MAP on arterial stiffness.

**Results:** Participants (n=5,942) were mean (standard deviation) age of 62.8 (10.3) years (52% female; 39% White, 25% African-American, 13% Asian, 23% Hispanic). Systolic BP (SBP) was 133.5 (20.5) mmHg, diastolic BP (DBP) was 74.27 (10.28) mmHg and pulse pressure (PP) was 59.22 (16.17) mmHg. Total carotid stiffness was associated with the SBP PRS (*β*=0.10 +/-0.02, p=5.25E-08) and the DBP PRS (*β*: 0.08 +/-0.02, p=7.26E-07) but not PP PRS (*β*=0.04 +/-0.02, P=0.05). Load dependent carotid stiffness was associated with the SBP PRS *β*=0.09 +/-0.01, p=7.93*10^-30^) the DBP PRS (*β*=0.09 +/-0.01, p= 4.92*10^-34^) and the PP PRS (*β*=0.03 +/-0.01, p= 8.18*^10-5^). Structural carotid stiffness was not associated with any BP PRSs. MR showed that SBP and DPB PRS strongly predict MAP (p<0.001) and each 1-mmHg increase in MAP raised load dependent stiffness by 0.0494 m/s (p<0.001).

**Conclusion:** There are key differences in the genetic underpinnings of the mechanistic components of arterial stiffness.

## Introduction

Arterial stiffness is a well-established risk-factor for cardiovascular disease (CVD) events and can enhance the accuracy of CVD risk assessments.^1-6^ The intricate relationship between normal aging, hypertension, and arterial stiffness is complex, as hypertension serves as both a causative factor and a consequence of worsening arterial stiffness.^2,7-13^ Recent investigations have proposed a novel framework where total arterial stiffness can be divided into two distinctive components: 1) structural arterial stiffness arising from arterial wall remodeling, and 2) load-dependent arterial stiffness associated with elevated blood pressure load on the artery wall.^14^ There is considerable heterogeneity between stiffness components among middle-aged and older adults at risk for CVD and both of these phenotypic traits have been identified as independent contributors to major adverse outcomes including the onset of hypertension.^14-16^ Together, this emphasizes the need to consider these two traits separately, particularly to identify disease mechanisms in arterial stiffness and new therapeutic targets.

While the heritability of blood pressure and hypertension are widely recognized, our understanding of the genetic underpinnings of arterial stiffness remains limited.^17-19^ As our comprehension of the genetic landscape contributing to hypertension deepens, it becomes imperative to explore the genetic impact of blood pressure on each distinct arterial stiffness trait. A recent genome wide analysis of over one million adults found 2,103 independent genetic signals identified for blood pressure traits offering a large amount of genetic information for BP and the generation of polygenic risk scores (PRS).^20^ Polygenic risk scores (PRSs), sum up the subtle effects of many alleles across the human genome, and offer a potential powerful tool for refining our ability to predict disease risk.

In this study we leverage previously identified blood pressure PRSs and conduct a comprehensive analysis to unravel the intricate associations between arterial stiffness traits and blood pressure PRSs, which will provide a deeper understanding of the relationships between arterial stiffness markers and variations in gene combinations. By identifying the genetic distinctions between total, structural, and load-dependent stiffness, we aim to provide novel insights into hypertension. This research endeavors to bridge the gap in our understanding, offering potential avenues for more personalized and effective approaches in the management of hypertension.

## Materials and Methods

The present analysis draws upon data from the Multi-Ethnic Study of Atherosclerosis (MESA), a prospective cohort comprising 6,814 participants recruited from six diverse communities across the United States.^21^ At baseline, MESA enrolled both men and women, aged 45-84 years, from four race/ethnic groups, who were free of overt cardiovascular disease (CVD). Additional exclusion criteria included: active treatment for cancer; pregnancy; any serious medical condition or cognitive inabilities which would prevent long-term participation; weight >300 pounds; plans to leave the community within five years; a lack of fluency in any of English/Spanish/Cantonese/Mandarin; and having undergone a chest CT scan in the past year. A detailed exposition of the study’s objectives and design has been previously published. Ethical considerations were paramount, with all participants providing informed consent for the study protocol, which received approval from the Institutional Review Boards of the ultrasound reading center and all MESA field centers.

All participants underwent an in-person clinic exam at baseline (2000-2002), during which a blood sample was taken. This analysis is based on a subset of MESA participants (n=6452) who underwent a valid carotid ultrasound examination (N=6028) at the first assessment and were not missing data needed to calculate arterial stiffness measures (N=5942). The demographic, medical history, and laboratory data used in this study was collected at the first examination conducted between July 2000 and August 2002.

### B-Mode Ultrasound and Brachial Blood Pressure Measurements

During examination 1, B-mode ultrasound video-loop recordings were captured from a longitudinal section of the distal right common carotid artery using a Logiq 700 ultrasound system (General Electric Medical Systems) with a transducer frequency of 13 MHz. These video images were meticulously digitized at high resolution and frame rates through a medical digital recording device (PACSGEAR, Pleasanton, CA), and subsequently converted into DICOM-compatible digital records.^22^ Trained and certified sonographers across all six MESA sites followed the same scanning conditions and imaging protocol. This involved maintaining consistent display depth, angle of approach, image optimization, and ultrasound system settings. Prior to ultrasound image acquisition, participants underwent a 10-minute rest in the supine position, during which repeated measures of brachial blood pressures were obtained using an automated upper arm sphygmomanometer (DINAMAP; GE Medical Systems, Milwaukee, WI). The obtained ultrasound images were meticulously reviewed and interpreted by the MESA Carotid Ultrasound Reading Center, located at the University of Wisconsin Atherosclerosis Imaging Research Program in Madison, WI. Systolic and diastolic diameters were ascertained as the largest and smallest diameters observed during the cardiac cycle. All measurements were conducted manually and in triplicate, tracing the last centimeter of the right carotid artery, spanning 2 to 3 consecutive cardiac cycles. Both internal and external artery diameters were precisely measured using Access Point Web version 8.2 (Freeland Systems, Westminster, CO).^22^

### Carotid Artery Stiffness Measurements

A detailed description of the common carotid artery stiffness calculations have been described previously and are included in the supplement.^14-16^ This includes calculation of total arterial stiffness, structural stiffness (determined using participant specific models and a reference blood pressure of 120/80 mmHg), and load dependent stiffness which is represented as the difference between total stiffness and structural stiffness.^14-16^

### Genome Wide Association data

Genomic DNA was isolated from whole blood for genetic analysis on all participants. Genome-wide data was obtained using the Affymetrix 6.0 chip and additional deeper sequencing using the Illumina HumanOmni chip. Exclusion criteria included heterozygosity > 53% and individual-level genotyping call rate < 95%. SNPs with call rate < 95% and monomorphic SNPs were removed. We further filtered on race/ethnic specific Hardy-Weinberg Equilibrium *p*-value > 1*10^-5^. The MiniMac4 program [PMID: 27571263] hosted on Michigan Imputation server (https://imputationserver.readthedocs.io/en/latest/) was used to impute GWAS data for MESA SHARe participants (chromosomes 1-22) to the 1000 Genomes reference panel (build GRChr37).

### Blood Pressure Polygenic Risk Scores

In this study, blood pressure polygenic risk scores (PRSs) for SBP DBP, and PP, were derived from genome-wide association studies (GWASs) involving over 1 million individuals with European-ancestry.^20^ The PRSs were reconstructed on individuals level data in MESA. A total of 2,103 blood pressure single nucleotide polymorphisms (SNPs) were included, with 1,780 in the SBP PRS, 1,761 in the DBP PRS, and 1,513 in the PP PRS. We included all SNPs showing associations with the respective trait in MESA that met a significance threshold of p<5×10-8. The weighted PRSs (PRS_SBP_, PRS_DBP_, PRS_PP_, and PRS_BP_) was calculated as:

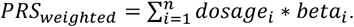

These PRSs were then used in association analyses within the four self-reported and genomically confirmed race and ancestry groups (European origin, African American, Hispanic, and Chinese), with standardization performed within each group before analysis.

### Covariates

Key health parameters were obtained at the baseline in-person clinic exam: Hypertension was defined as SBP ≥140 mm Hg, DBP ≥90 mm Hg, or the use of antihypertensive medications. Diabetes mellitus was identified by fasting blood glucose levels (12-hour fast) ≥126 mg/dL or the use of antiglycemic medications. Additionally, impaired fasting glucose was delineated as blood glucose ranging from 100 to 125 mg/dL. Lipid profiles, encompassing total and high-density lipoprotein cholesterol, were assessed after a 12-hour fast, with low-density lipoprotein cholesterol being calculated.^21^

### Study Subjects

MESA study included sample from four different race and ethnicity groups. Race and ethnicity was self-reported and ancestry was genomically confirmed using GWAS data. We carried out principle component analysis using SMARTPCA (Patterson et al., 2006; Price et al., 2006) within EIGENSTRAT and population outliers were identified as more than 5 standard deviation from the mean. Based on this procedure, 42 individuals from the Caucasian group, 19 from the Chinese American group, 7 from the African American group, and 70 from the Hispanic group were identified as population outliers and were removed from further analyses.

### Statistical Analyses

All analyses were conducted using SAS software (version 9.4). Distributions of all covariates were examined using measures of central tendency and a visual inspection of histograms. Normality was defined as having both a skew and kurtosis in the range of -1 to +1. Where distributions did not meet these criteria, variables were transformed to normality using Log-transformations or inverse normal transformations. No normal transformation was needed for the Carotid Artery Stiffness measurements.

#### Participant Characteristics

Raw (untransformed) data were used to provide means (+/-standard deviation; SD) for continuous variables, or total number (N) and percentage (%) for categorical or ordinal variables, stratified by self-reported and genomically confirmed race/ancestry and sex. Differences in demographic and health information between sex- and race/ancestry groups were examined using linear regression models for continuous variables and chi-squared tests of difference for categorical variables. For these descriptive analyses, P<.05 was retained for significance (i.e., without a correction for multiple testing).

#### PRS-phenotype Associations

The associations between baseline total stiffness, load-dependent stiffness, and structural stiffness with PRSs were examined using a multivariable-adjusted regression model, accounting for age, sex, body mass index (BMI), and study site. First, the association of each PRS (N=3) with each phenotype (N=3) in 9 regression models. Subsequently, the same models were rerun, stratified by gender (N=18 models), and then by race/ancestry (N=36 models). Significance was set at a Bonferroni corrected P<7.9*10^-4^ (.05/(9+18+36).

Mendelian Randomization (MR) was performed using the two-stage least squares method for each PRS (instruments). Load-dependent stiffness as the outcome variable, MAP as the exposure variable with age, sex and ancestry as covariates. The first stage used the PRSs and covariates to predict MAP and the second stage estimated the effect of MAP on LD. Weak instruments, Wu-Hausman and Sargan tests were used to determine reliability and support causal inference.^23-25^

#### Sensitivity analysis

To evaluate overlap between SNPs, we also performed multivariable models evaluating only SNPs from the SBP PRS (1419 SNPs), only SNPs from DBP (1400 SNPs) and SNPs that overlapped between SBP PRS and DBP PRS (361 SNPs). Additional models were performed which included baseline SBP in the model. Analyses were conducted within each race/ancestry group and the combined sample, with additional adjustment for race/ancestry in the latter.

## Results

### Baseline Participant Characteristics

The baseline participant population for the MESA cohort consisted of 6,814 men and women, aged 45 to 84 years, with self-reported White (N=2,623), Black (N=1,891), Hispanic (N=1,496), and Chinese (N=804). The current analyses included 5,804 participants with genomically confirmed race/ancestry data and arterial stiffness data. Baseline participant characteristics, stratified by gender, are detailed in **Table 1**. At baseline, participants had a mean age of 62.8 (10.3) years, and 52% of the participants were female. The racial distribution of the cohort was as follows: 39% white, 25% black, 13% Chinese, and 23% Hispanic. The average baseline blood pressure measurements were 133.5 (20.5) mmHg for systolic, 74.27 (10.28) mmHg for diastolic, and 59.22 (16.17) mmHg for pulse pressure. Total carotid stiffness, calculated as pulse wave velocity (cPWV), averaged 7.19 (1.65) m/s at baseline and structural stiffness and load-dependent stiffness PWV were of 7.18 (1.44) m/s and 0.01 (0.61) m/s, respectively (**able 1**). Values of stiffness parameters by sex and race/ancestry and differences are shown in **supplemental table 1 and table 1A**.

**Table 1:** Baseline Characteristics by Ancestry and Sex.

|  | ALL | African American<br>(n=1476) |  | Chinese<br>(n=727) |  | European American<br>(n=2298) |  | Hispanic<br>(n=1303) |  | p-value for differences |  |
| --- | --- | --- | --- | --- | --- | --- | --- | --- | --- | --- | --- |
|  |  | Male | Female | Male | Female | Male | Female | Male | Female | Race/<br>Ancestry* | Gender** |
| N | 5804 | 677 | 799 | 357 | 370 | 1111 | 1187 | 646 | 657 | - | - |
| Age (years) | 62.8<br>(10.3) | 63.2<br>(10.3) | 62.8<br>(10.1) | 63.1<br>(10.2) | 62.9<br>(10.4) | 63.3<br>(10.2) | 63<br>(10.4) | 61.6<br>(10.3) | 62.3<br>(10.3) | 0.01 | 0.78 |
| BMI (kg/m<sup>2</sup>) | 28.2<br>(5.4) | 31.2<br>(6.4) | 28.5<br>(4.6) | 23.8<br>(3.4) | 24.1<br>(3.2) | 27.4<br>(5.7) | 27.9<br>(4.0) | 30<br>(5.5) | 28.8<br>(4.3) | <.0001 | <.0001 |
| SBP (mmHg) | 133.5<br>(20.5) | 139<br>(18.4) | 139.3<br>(22.4) | 129.8<br>(19.1) | 131.1<br>(22.9) | 132.1<br>(17.0) | 129.1<br>(20.3) | 133.5<br>(19.9) | 134.6<br>(22.7) | <.0001 | 0.30 |
| DBP (mmHg) | 74.3<br>(10.3) | 80.4<br>(9.4) | 75<br>(11.0) | 77.5<br>(9.4) | 70.7<br>(10.3) | 76.8<br>(8.6) | 69.3<br>(9.5) | 77<br>(8.9) | 69.4<br>(9.4) | <.0001 | <.0001 |
| PP (mmHg) | 59.3<br>(16.1) | 58.6<br>(14.3) | 64.3<br>(17.4) | 52.4<br>(14.7) | 60.4<br>(17.8) | 55.3<br>(12.8) | 59.8<br>(15.9) | 56.5<br>(15.8) | 65.2<br>(18.7) | <.0001 | <.0001 |
| HTN status | 2557<br>(44.1%) | 384<br>(56.7%) | 489<br>(61.2%) | 125<br>(35.0%) | 146<br>(39.5%) | 434<br>(39.1%) | 446<br>(37.6%) | 243<br>(37.6%) | 290<br>(44.1%) | <.0001 | <.0001 |
| Anti-HTN medication use | 2119<br>(36.5%) | 319<br>(47.1%) | 416<br>(52.2%) | 98<br>(27.5%) | 107<br>(28.9%) | 380<br>(34.2%) | 380<br>(32.0%) | 195<br>(30.1%) | 224<br>(34.1%) | <.0001 | <.0001 |
| Total Stiffness (m/s) | 7.19<br>(1.7) | 7.5<br>(1.8) | 7.5<br>(1.8) | 7.2<br>(1.5) | 7.4<br>(1.8) | 6.9<br>(1.4) | 6.9<br>(1.5) | 7.2<br>(1.6) | 7.4<br>(1.8) | <.0001 | 0.009 |
| Structural Stiffness (m/s) | 7.18<br>(0.14) | 7.2<br>(1.7) | 7.4<br>(1.6) | 7.1<br>(1.4) | 7.5<br>(1.5) | 6.8<br>(1.2) | 7.2<br>(1.3) | 7.1<br>(1.3) | 7.6<br>(1.5) | <.0001 | <.0001 |
| Load-Dep. Stiffness (m/s) | 0.01<br>(0.6) | 0.3<br>(0.5) | 0.1<br>(0.7) | 0.1<br>(0.6) | -0.1<br>(0.7) | 0.1<br>(0.5) | -0.2<br>(0.6) | 0.1<br>(0.6) | -0.1<br>(0.6) | <.0001 | <.0001 |
| SBP PRS | 465.5<br>(6.5) | 466.8<br>(5.8) | 466.5<br>(5.8) | 466.5<br>(5.7) | 466.4<br>(5.4) | 464.1<br>(7.0) | 464.3<br>(7.0) | 466<br>(6.5) | 465.8<br>(6.7) | <.0001 | 0.87 |
| DBP PRS | 288.8<br>(4.1) | 288.8<br>(3.5) | 288.6<br>(3.6) | 290.8<br>(3.0) | 290.4<br>(3.2) | 288.6<br>(4.7) | 288.7<br>(4.4) | 288.5<br>(4.1) | 288.2<br>(4.2) | <.0001 | 0.19 |

\*Differences by genetically defined ancestry across both genders
\*\*Differences by gender across all genetically defined ancestry groups
Abbreviations: BMI: body mass index; SBP: Systolic blood pressure; DBP: Diastolic blood pressure; PP: Pulse pressure; HTN: Hypertensive; Load-Dep.: Load-dependent; PRS: Polygenic risk score.

### Blood Pressure Polygenic Risk Scores with Systolic and Diastolic Blood Pressure and Pulse Pressure

Blood pressure PRS distributions by race/ancestry are shown in **Figure 1**. Associations between the SBP, DBP, and PP traits in MESA were strongly associated with the corresponding blood pressure PRSs (all p > 4.96*10^-34^, **Supplemental Table 2**).

**Figure 1:**
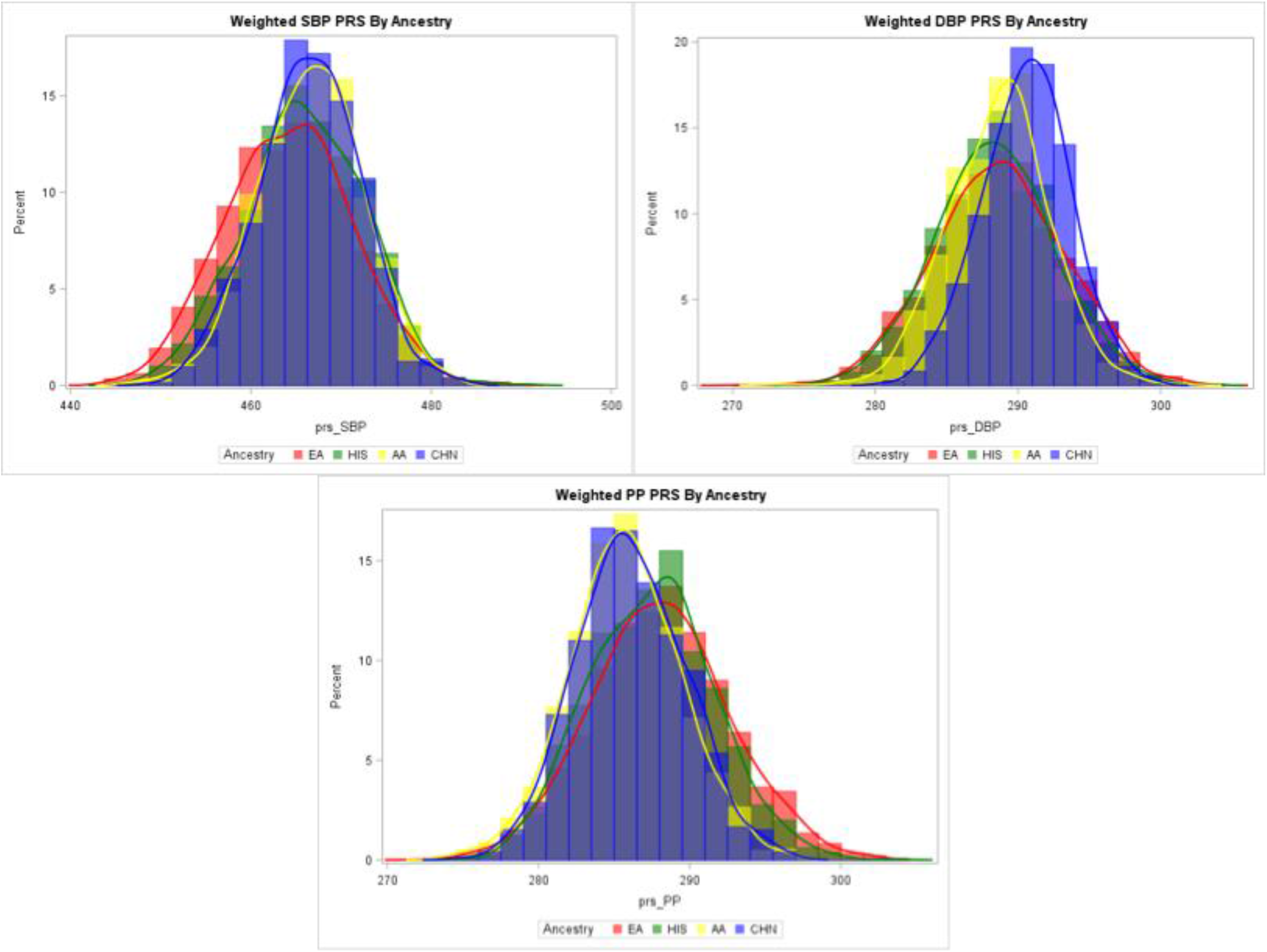
Blood Pressure PRS Distributions by Race/Ancestry.

Associations between SBP, DBP and PP PRSs and baseline carotid artery stiffness mechanisms for all participants are shown in **Table 2**. In the unadjusted models, total carotid stiffness was associated with the SBP PRS (*β*: 0.10 +/-0.02, p=5.25E-08) and the DBP PRS (*β*: 0.08 +/-0.02, p=7.26E-07) but not PP PRS (*β*=0.04 +/-0.02, P=0.05). Load dependent carotid stiffness was also associated with the SBP PRS *β*=0.09 +/-0.01, p=7.93*10^-30^) the DBP PRS (*β*=0.09 +/ -0.01, p= 4.92*10^-34^) and the PP PRS (*β*=0.03 +/-0.01, p= 8.18*^10-5^). Structural carotid stiffness was not associated with any of the three BP PRSs (all p>.05; **Table 2**).

**Table 2:** Standardized Parameter Estimates for the Associations between Blood Pressure Polygenic Risk Scores and Carotid Artery Stiffness Mechanisms in the MESA Population.

|  | SBP PRS |  | DBP PRS |  | PP PRS |  |
| --- | --- | --- | --- | --- | --- | --- |
| | $\beta$(SE) | P | $\beta$(SE) | P | $\beta$(SE) | P |
| Total Carotid Stiffness | **0.10(0.02)** | **5.25\*10<sup>-8</sup>** | **0.08 (0.02)** | **7.26\*10<sup>-7</sup>** | 0.04(0.02) | .05 |
| Structural Carotid Stiffness | 0.02(0.02) | .29 | 0.002(0.02) | .89 | 0.01(0.02) | .67 |
| Load-Dependent Carotid Stiffness | **0.09(0.01)** | **7.93\*10<sup>-30</sup>** | **0.09 (0.01)** | **4.92\*10<sup>-34</sup>** | **0.03(0.01)** | **8.18\*10<sup>-5</sup>** |

*Note:* All models adjusted for age, sex, ancestry, BMI and study site. Bold indicates significance at a Bonferroni corrected P<0.001 *Abbreviations:* BMI: body mass index; SBP: Systolic blood pressure; DBP: Diastolic blood pressure; PP: Pulse pressure; PRS: Polygenic risk score.

When stratified by sex, similar associations were noted, with the exception that the association of total carotid stiffness with DBP was not significant in males (*β*: 0.06 +/-0.03, p=0.02), although it remained so in females (*β*: 0.12 +/-0.02, p=4.62E-06, Table 3). Load dependent carotid stiffness remained significantly associated with both PRSs in both male and females (**Table 3**). Load dependent stiffness was also associated with the PP PRS only in males (*β*: 0.03 +/ - 0.01], p=0.001; Table 3). Once again, there were no associations with structural carotid stiffness and any of the three BP PRSs or PP PRS (P>.05; Table 3).

**Table 3:**
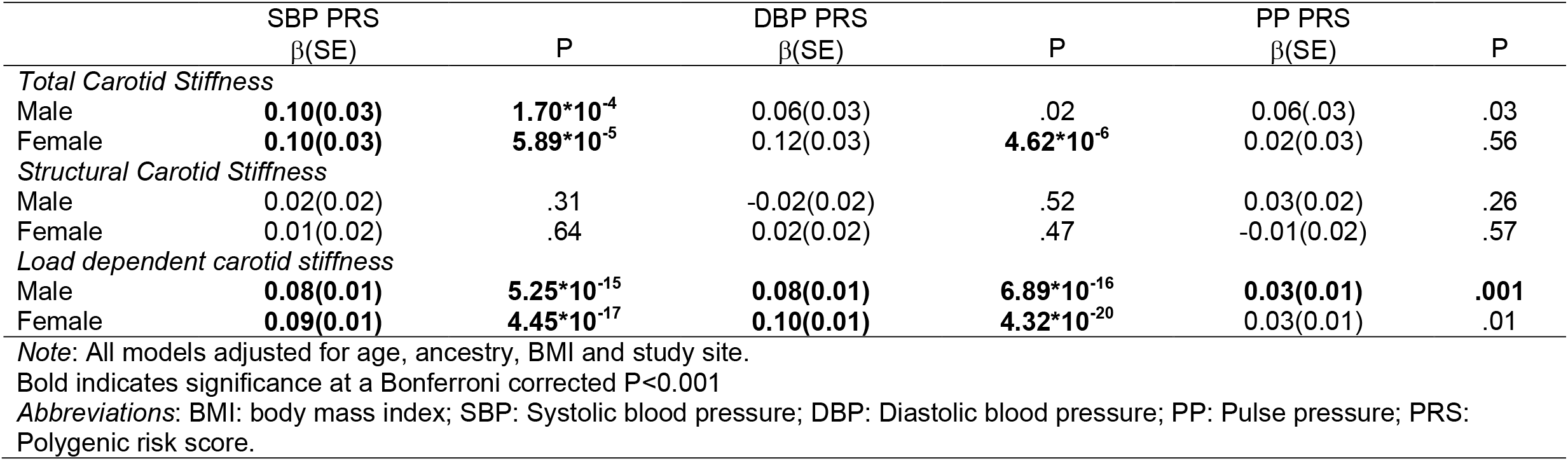
Standardized Parameter Estimates for the Associations between Blood Pressure Polygenic Risk Scores and Carotid Artery Stiffness Mechanisms in the MESA Population, stratified by sex.

**Table 4** shows associations between the BP PRSs and stiffness measures by race/ancestry. Notably, there were very weak associations with total carotid stiffness and SBP PRS for participants with European American (EA) ancestry (*β*: 0.09 +/ - 0.03, p=0.0004), but no other race/ancestry groups. Conversely, total stiffness was weakly associated with DBP PRS in those with Hispanic (*β*: 0.15 +/-0.04, p=0.0001). Load-dependent stiffness had strong and significant associations with SBP and DBP PRSs for those with EA (all p≤ 7.48E-19), Hispanic (all p≤ 4.45E-15) and Chinese (all p≤ 2.36E-05). There were no significant associations between SBP PRS and DBP PRS in those with AA. There were no significant associations with PP PRS in any race/ancestry group.

**Table 4:** Associations between Blood Pressure Polygenic Risk Scores and Carotid Artery Stiffness Mechanisms in the MESA Population, stratified by genetically-defined ancestry groups.

| | SBP PRS<br>$\beta$(SE) | P | DBP PRS<br>$\beta$(SE) | P | PP PRS<br>$\beta$(SE) | P |
| --- | --- | --- | --- | --- | --- | --- |
| *Total Carotid Stiffness* |  |  |  |  |  |  |
| African American | 0.09(0.04) | .03 | 0.10(0.04) | .02 | -0.004(0.04) | .93 |
| Chinese | 0.10(0.05) | .06 | 0.03(0.05) | .61 | 0.12(0.05) | .03 |
| European American | **0.09(0.03)** | **4.0\*10<sup>-4</sup>** | 0.07(0.03) | .01 | 0.06(0.03) | .03 |
| Hispanic | **0.13(0.04)** | **.001** | **0.15(0.04)** | **1.0\*10<sup>-4</sup>** | **0.003(0.04)** | **.001** |
| *Structural Carotid Stiffness* |  |  |  |  |  |  |
| African American | 0.06(0.04) | .13 | 0.06(0.04) | .10 | 0.0004(0.04) | .96 |
| Chinese | 0.002(0.05) | .96 | -0.07(0.05) | .12 | 0.05(0.05) | .26 |
| European American | -0.003(0.03) | .91 | -0.03(0.02) | .91 | 0.02(0.02) | .40 |
| Hispanic | 0.01(0.03) | .79 | 0.02(0.03) | .54 | -0.03(0.03) | .32 |
| *Load dependent carotid stiffness* |  |  |  |  |  |  |
| African American | 0.03(0.02) | .03 | 0.04(0.02) | .02 | -0.004(0.02) | .78 |
| Chinese | **0.09(0.02)** | **2.36\*10<sup>-5</sup>** | **0.10(0.02)** | **5.63\*10<sup>-6</sup>** | 0.06(0.02) | .004 |
| European American | **0.10(0.01)** | **7.48\*10<sup>-19</sup>** | **0.10(0.01)** | **1.48\*10<sup>-20</sup>** | **0.04(0.02)** | **7.0\*10<sup>-4</sup>** |
| Hispanic | **0.12(0.01)** | **4.45\*10<sup>-15</sup>** | **0.13(0.01)** | **1.25\*10<sup>-15</sup>** | 0.04(0.01) | .02 |

Note: Ancestry reference is European Ancestry
Note: All models adjusted for age, sex, BMI and study site.
Bold indicates significance at a Bonferroni corrected P<0.001
Abbreviations: BMI: body mass index; SBP: Systolic blood pressure; DBP: Diastolic blood pressure; PP: Pulse pressure; PRS: Polygenic risk score.

Associations with carotid stiffness measures are shown in **Table 5**. When age, sex and race/ancestry were included in multivariate models, the strongest associations were seen with load dependent stiffness and the SBP PRS (*β*: 0.05 +/-0.01, p<0.0001 and DBP PRS (*β*: 0.06 +/-0.01], p<0.0001), with weaker associations with total carotid stiffness and SBP PRS (*β*: 0.07 +/-0.02, p=0.003) and DBP PRS (*β*: 0.05 +/-0.02, p=0.04). There were no significant associations with structural carotid stiffness and SBP or DBP PRS (all p>0.22). When baseline SBP was included in the models, load dependent carotid stiffness remained associated with SBP PRS and DBP PRS (p<0.001), but total stiffness and structural stiffness were not (all p>0.2, **supplemental table 3**). To further understand how overlapping SNPs were impacting the associations, we performed a sensitivity analysis by creating three additional PRSs (**supplemental table 4**): 1) SNPs that were only included in the SBP PRS (1419 SNPs), 2) SNPs that were only included in the DBP PRS (1400 SNPs) and 3) SNPs that were in both the SBP and DBP PRS (361 SNPs). Only load-dependent carotid stiffness was associated with SBP only SNPs (*β*: 0.02 +/-0.01, p=0.006), total stiffness and structural stiffness were not (p>0.3). In the PRS with overlapping SNPs, there were moderate negative associations with total stiffness (*β*: -0.05 +/-0.02, p=0.01) and load dependent stiffness (*β*: -0.02 +/-0.01, p=0.04) but not structural stiffness (*β*: -0.03 +/-0.02, p=0.06).

**Table 5.** Multivariate Models for Stiffness Mechanisms with SBP and DPB PRSs, Age, Sex and Ancestry.

|  | Total Carotid Stiffness |  |  | Structural Carotid Stiffness |  |  | Load-Dep. Carotid Stiffness |  |  |
| --- | --- | --- | --- | --- | --- | --- | --- | --- | --- |
| | $\beta$ | SE | P | $\beta$(SE) | SE | P | $\beta$(SE) | SE | P |
| SBP PRS | **0.07** | **0.02** | **0.0025** | 0.03 | 0.02 | 0.2218 | **0.05** | **0.01** | **<.0001** |
| DBP PRS | 0.05 | 0.02 | 0.0431 | -0.01 | 0.02 | 0.5143 | **0.06** | **0.01** | **<.0001** |
| Age | **0.08** | **0.00** | **<.0001** | **0.06** | **0.00** | **<.0001** | **0.01** | **0.00** | **<.0001** |
| Male Sex | **-0.11** | **0.04** | **0.002** | **-0.35** | **0.03** | **<.0001** | **0.24** | **0.01** | **<.0001** |
| African American | **0.60** | **0.05** | **<.0001** | **0.30** | **0.04** | **<.0001** | **0.30** | **0.02** | **<.0001** |
| Chinese | **0.38** | **0.06** | **<.0001** | **0.34** | **0.05** | **<.0001** | 0.04 | 0.02 | 0.0779 |
| Hispanic | **0.52** | **0.05** | **<.0001** | **0.43** | **0.04** | **<.0001** | **0.09** | **0.02** | **<.0001** |

*Note:* Ancestry reference is European Ancestry
*Note:* All models adjusted for age, sex, BMI and study site.
Bold indicates significance at a $P < 0.01$
*Abbreviations:* BMI: body mass index; SBP: Systolic blood pressure; DBP: Diastolic blood pressure; PP: Pulse pressure; PRS: Polygenic risk score.

Mendelian Randomization showed that, in the first stage, both the SBP PRS (β = 0.94, p < 0.001) and DBP (β = 1.22, p < 0.001) strongly predict MAP (F-statistic = 160.9 [p < 0.001], R^2^ = 0.1547). The second stage demonstrated that MAP had a causal effect on load-dependent stiffness (β = 0.0494, p < 0.001). Each 1-mmHg MAP increase raises load-dependent stiffness by 0.0494 m/s, showing a significant positive causal effect of MAP on load-dependent stiffness. The instruments were found to be reliable and supported causal inference (Weak Instruments: F = 89.88, p < 0.001 [strong instruments]; Wu-Hausman: p = 0.219 [no strong endogeneity]; Sargan: p = 0.419 [instruments valid].^23-25^

## Discussion

We evaluated associations between novel arterial stiffness traits (including total arterial stiffness, structural arterial stiffness and load-dependent arterial stiffness) and polygenic risk scores for blood pressure parameters including SBP, DBP, and PP. Our findings revealed robust and consistent associations between load-dependent stiffness and both systolic blood pressure polygenic risk scores (SBP PRS) and diastolic blood pressure polygenic risk scores (DBP PRS), even after adjusting for age, race/ancestry, and gender. In contrast, associations between total stiffness and SBP PRS are much weaker and there were no significant associations with structural stiffness and any BP PRSs. Lastly, we found that MAP causally increases load-dependent stiffness using mendelian randomization (MR).

These finding holds clinical significance as load-dependent stiffness emerges as a promising biomarker for hypertension risk assessment and management, highlighting its potential as a meaningful marker of CVD risk beyond traditionally measured total stiffness (where associations are likely diluted by elements of structural stiffness) and blood pressure alone. Bridging the functional gap between the identified SNPs/PRSs and the phenotypic traits of arterial stiffness is an essential launching point to improving risk prediction and blood pressure care in adults through precision medicine and personalized medicine.^26,27^ Prior studies to evaluate associations between SNPs and arterial stiffness had smaller sample sizes^28-30^ and used far fewer SNPs^28,29,31,32^. Though the PRSs were specific to blood pressure, it has been shown that increasing the number of SNPs in genetic risk scores can significantly improve risk prediction.^33^ Moreover, prior studies attempting to identify SNPs associated with total arterial stiffness have yielded inconsistent results, particularly when these studies have been performed in ethnically different ancestry groups.^30,32^

A previous study by Cecelja et al. identified 896 SNPs that were robustly associated with blood pressure and applied them to total stiffness measured by carotid to femoral PWV.^19^ They also found subtle but measurable differences between the SNPs associated with SBP compared to DBP and total arterial stiffness. They also found that total arterial stiffness had the highest heritability, with the greatest number of genes shared between SBP and arterial stifness.^19^

The drastic differences between associations with load-dependent stiffness and structural stiffness with blood pressure PRSs supports our hypothesis that these metrics are genetically distinct. Load-dependent stiffness is primarily influenced by the mechanical forces exerted on the arterial wall due to acute changes in blood pressure. Genetic factors affecting blood pressure regulation^19,26^, captured by the polygenic risk scores, may more directly impact the loading mechanics of the arterial wall, leading to a stronger association with load-dependent stiffness. Arterial stiffness has been shown to be strongly heritable,^19^ and genetic variants related to blood pressure regulation could be more effectively captured with load-dependent stiffness.

Conversely, the intrinsic changes in the blood vessel wall that comprise structural stiffness is predominantly due to the structural changes in the arterial wall that occur with aging and was strategically developed to exclude the component of arterial stiffness that is acutely related to the distending effect of blood pressure.^14,34^ Alterations in the structural stiffness of large elastic arteries is predominantly impacted by changing the composition of collagen, elastin and fibrin.^2^ Studies in spontaneously hypertensive rats also suggests that vascular smooth muscle cells play a role in the structural stiffening of arteries that occurs with aging.^35^ Our finding that blood pressure PRSs are not associated with increased structural artery stiffness provides novel support to past findings that hypertension and blood pressure do not increase structural arterial stiffness in middle-aged and older adults. ^15,36-39^ Genetic factors influencing structural stiffness may be more complex and multifaceted, involving a broader array of molecular and cellular processes.^40,41^ As a result, the polygenic risk scores derived from blood pressure-related genetic variants may not comprehensively represent the genetic complexity underlying structural stiffness. The changes in structural components of the arterial wall occur over longer periods of time^15^ and may be influenced by a combination of genetic and environmental factors, or could be more closely linked with the genetic determinants of cholesterol metabolism and atherosclerosis, which would not be fully captured by the polygenic risk scores derived from blood pressure-related SNPs.

We also observed small, yet interesting differences between, DBP and SBP PRSs. Diastole represents the phase of the cardiac cycle when the heart relaxes, and blood is refilled into the coronary arteries and systemic circulation.^2,9,42,43^ SBP is a pulsatile component of blood pressure that is directly related to stroke volume and arterial stiffness, while DBP makes up a much greater proportion of MAP (2/3) and is determined by cardiac output and systemic vascular resistance.^19^ Genetic factors that affect diastolic blood pressure regulation may play a more direct role in determining the compliance of arterial walls. While SBP PRS may capture the genetic predisposition to elevated systolic pressure, the pulsatile nature of systolic stress might not be as directly linked to changes in arterial stiffness compared to the continuous stress experienced during diastole. Further research is warranted to unravel the intricate mechanobiologic pathways underlying these associations and their clinical implications.

Understanding the differential genetic impact on arterial stiffness components has profound clinical implications, specifically being able to identify individuals at risk prior to the development of CVD. Load-dependent stiffness continues to emerge as a promising phenotypic trait that could serve as a sensitive marker for cardiovascular risk, aiding in risk stratification and targeted interventions, both at the phenotypic and genotypic level. The observed divergence between structural stiffness and blood pressure genetics also opens new avenues for therapeutic strategies focusing on the intrinsic properties of the blood vessels that are distinct from blood pressure control and current antihypertensive medications.

While there were similarities between those with Chinese, EA, and Hispanic, we did not find any significant associations between the BP PRSs with load-dependent stiffness in African Americans. This could be due to the fact that most of the samples used to determine the blood pressure PRSs were acquired in those with genetic ancestry that is predominantly EA and ancestry specific SNPs (related to different disease mechanisms) that could drive this association are not represented in our current PRS. Alternatively, it could be that elevated load-dependent stiffness in individuals with African American is driven by social determinants of health instead of genetics. Limited access to healthcare and substandard insurance coverage have previously been shown as key factors contributing to racial disparities in BP control^44^ and/or gene life-style interactions that need to be better captured Additionally, it may indicate that the BP and load-dependent stiffness in African Americans has a different mechanism, or simply indicate that because the genetic effect size in African American was smaller and our limited sample size in African Americans did not have enough power to detect the association at the level of significance. This has direct clinical implications, specifically since it is known that certain antihypertensive medications classes have improved outcomes in African American individuals.^45,46^

These disparities underscore the importance of considering genetic determinants within specific demographic subgroups when assessing cardiovascular risk. The observed pattern suggests that the genetic determinants of arterial stiffness are complex and multifactorial. Load-dependent mechanisms, being more directly influenced by acute changes in blood pressure, may exhibit stronger associations with blood pressure PRS, while the genetic regulation of structural stiffness may involve a broader and more intricate set of factors not fully captured by the examined polygenic risk scores of blood pressure. Further research using a comprehensive approach that explore muti-trait PRSs for other potential risk factors (atherosclerosis, peripheral artery disease, cholesterol, aortic aneurysms, etc) is warranted to dissect and inform the specific genetic contributors to these distinct components of arterial stiffness. Additionally, subgroup analysis and gene life-interaction in high risk groups for CV outcomes may have a magnify benefit on risk prediction.

### Limitations and Future Directions

While our study advances understanding, it is not without limitations. Our findings emphasize the need for further exploration of the specific genetic loci contributing to directly to the stiffness mechanisms, not just blood pressure. We also recognize that the PRSs used were constructed based on blood pressure phenotypes from predominantly EA ancestry groups, thus SNPs related to arterial stiffness metrics that are not directly related to blood pressure or ancestry groups that are dissimilar from EA will require further study. Furthermore, ideally central blood pressure would be used to calculate arterial stiffness measures; however this study used peripheral blood pressure as a surrogate for central blood pressure, as central blood pressure was not measured at exam 1. While we did perform Mendelian randomization, reverse causation (load dependent stiffness to MAP) could not be tested due to lack of known relationships between SNPs and the arterial stiffness components.

## Conclusions

In conclusion, our study underscores that there are different genetic mechanisms underscoring structural and load dependent arterial stiffness. We demonstrate that a major genetic determinant of load-dependent arterial stiffness are the genes responsible for variation in systolic blood pressure. The observed disparities in genetic associations across demographic subgroups may also be used to help personalize our approach to understanding and managing arterial stiffness. As we move toward an era of precision medicine, our findings contribute crucial insights into the intersection of genetics and cardiovascular health, opening avenues for targeted interventions based on individualized risk profiles.

## Data Availability

Data is available through the MESA website.

https://mesa-nhlbi.org/

## Acknowledgements

MESA and the MESA SHARe projects are conducted and supported by the National Heart, Lung, and Blood Institute (NHLBI) in collaboration with MESA investigators. Support for MESA is provided by contracts 75N92020D00001, HHSN268201500003I, N01-HC-95159, 75N92020D00005, N01-HC-95160, 75N92020D00002, N01-HC-95161, 75N92020D00003, N01-HC-95162, 75N92020D00006, N01-HC-95163, 75N92020D00004, N01-HC-95164, 75N92020D00007, N01-HC-95165, N01-HC-95166, N01-HC-95167, N01-HC-95168, N01-HC-95169, UL1-TR-000040, UL1-TR-001079, and UL1-TR-001420, UL1TR001881, DK063491, and R01HL105756. Funding for SHARe genotyping was provided by NHLBI Contract N02-HL-64278. Genotyping was performed at Affymetrix (Santa Clara, California, USA) and the Broad Institute of Harvard and MIT (Boston, Massachusetts, USA) using the Affymetrix Genome-Wide Human SNP Array 6.0. The authors thank the other investigators, the staff, and the participants of the MESA study for their valuable contributions. A full list of participating MESA investigators and institutes can be found at http://www.mesa-nhlbi.org. Adriana M. Hung is supported by a VA CSR&D MVP grant for the Genetic of Kidney Disease and Hypertension (CX001897).

